# Evaluation of viral loads in patients with SARS-CoV-2 Delta variant infection: Higher loads do not translate into different testing scenarios

**DOI:** 10.1101/2021.10.14.21265031

**Authors:** Juan Luis Gomez Marti, Ashley Mays, Melissa McCullough, Alan Wells, Tung Phan

## Abstract

The Delta SARS-CoV-2 variant is very infectious, and it is spreading quickly during this pandemic. In the study, we compared viral loads in surging cases infected with the SARS-CoV-2 Delta variant in the fourth wave of COVID-19 with the three prior waves. The data comprised viral loads from positive cases detected within the UPMC health care system in Allegheny County, Pennsylvania. A total of 2,059 upper airway samples were collected and tested for SARS-CoV-2 positive by RT-PCR during March 2020-September 2021. We did not observe significant difference in viral load difference between the third (December 2020 – January 2021) and fourth (June 2021 – September 2021) waves; however, they had the higher viral load than the first (March 2020 – June 2020) and second waves (June 2020 – August 2020). We did find an age-related effect with the elderly presenting with lower viral loads, which was also seen in the earlier waves. However, the level of viral load in the fourth wave was not sufficient higher to qualitatively change our expected detected rates using various testing modalities.

## The study

Coronaviruses are enveloped positive-stranded RNA viruses, and they belong to the genus *Coronavirus* of the family *Coronaviridae*. They are the single largest group of viruses, which has been associated with a variety of diseases in humans and animals (1). One of them is the Severe Acute Respiratory Syndrome Coronavirus 2 (SARS-CoV-2), which is responsible for the coronavirus disease 2019 (COVID-19) pandemic in humans (2). This virus causes a wide range of signs and symptoms from a mild condition to a very severe illness that requires hospitalization and intensive care (3). Transmission of SARS-CoV-2 is primarily by the respiratory route, which can happen directly from person to person (4). There is strong evidence that SARS-CoV-2 can spread by airborne transmission (5). The virus has been detected not only in respiratory specimens (nasopharynx, nose, bronchoalveolar lavage, sputum, and saliva) but also in non-respiratory specimens (feces, blood, and cerebral spinal fluid) (6, 7). Even though several COVID-19 vaccines are licensed globally, SARS-CoV-2 infection is still a significant public health concern. The United States is one of the emerged epicenters of the COVID-19 pandemic having more than 44 million confirmed cases and at least 700,000 deaths as of October 10^th^, 2021 (https://coronavirus.jhu.edu/map.html). Thus, testing to detect SARS-CoV-2 virus remains critical to diagnosing patients and handling the pandemic.

Every virus mutates, and there is nothing exceptional about SARS-CoV-2 in the context of evolution. What we have all witnessed is SARS-CoV-2 continuing to mutate rapidly worldwide, with each new variant more infectious than the last. The highly transmissible Delta variant (B.1.617.2) was first reported in India in December 2020 (https://gvn.org/covid-19/delta-b-1-617-2/), and it quickly became the predominant strain in the United States. There are reports that the reason for the Delta virus replacing all other variants in the United States by the summer of 2021, in addition to greater infectivity, is that the Delta variant presents with much higher viral loads in the upper airways. This would have implications for the testing modalities, as various rapid antigen tests require significantly higher viral loads for detection of infection (8).

Herein, we provide a real-world experience study in which evaluated viral loads in the Delta variant infections in the 4^th^ wave of COVID-19 and compared that with our data from the three prior waves. To be consistent in the population base, we queried the comprehensive clinical data of those persons presenting to the UPMC health care system hospitals in Allegheny County, Pennsylvania.

Between March 2020 and September 2021, a total of 2,059 nasopharyngeal specimens (459, 163, 988, and 449 in the 1^st^-4^th^ waves of COVID-19, respectively) were collected from symptomatic patients and tested as SARS-CoV-2 positive by the NAAT assays (Cepheid GeneXpert Xpress SARS-CoV-2 assay or and LDT based upon the CDC 2019-nCoV Real-Time RT-PCR Diagnostic Panel). These NAAT assays were performed as part of routine medical care, and they have been authorized by FDA under an Emergency Use Authorization (EUA). The cycle threshold (Ct) values generated by the two NAAT assays are not identical but have been shown to be relatively comparable (9). Viral loads were estimated by the Ct values. Statistical analyses were performed using with the Kruskal-Wallis test to compare more than two groups.

As shown in Fig. 1a, there was no significant difference between the viral loads of the 3^rd^ (median Ct 22.0) and 4^th^ (median Ct 23.0) waves. However, these Ct values are much lower (meaning higher viral load) than those of the 1^st^ (median Ct 26.6) and 2^nd^ (median Ct 26.5) waves. The viral load of the Delta variant on the patients by different age groups (15-24, 25-49, 50-64, 65-74, and ≥ 75 years) was evaluated in the 4^th^ wave. The median Ct value of the Delta variant RNA ranged from 21.7 to 28.0 depending on the age groups (Fig. 2). Of note, the viral load was highest in the age groups of 25-49 years (median Ct 22.3) and 50-64 (median Ct 21.7) years, followed by the age groups of 15-24 years (median Ct 23.8) and 65-74 (median Ct 23.6) years. The patients aged ≥75 years had the median Ct 28.0. Compared to the previous waves, the Delta variant was found to be associated with the increased viral load in the patients aged 25-74 years as shown in Fig. 1b. In each age group of 25-49, 50-64, and 65-74 years, the viral loads (median Ct 22.3, 21.7, and 23.6) in the 4^th^ wave was higher than those in the 1^st^ wave (median Ct 26.9, 26.7, and 27.9) and 2^nd^ wave (median Ct 25.9, 24.4, and 26.4).

**Figure 1.**
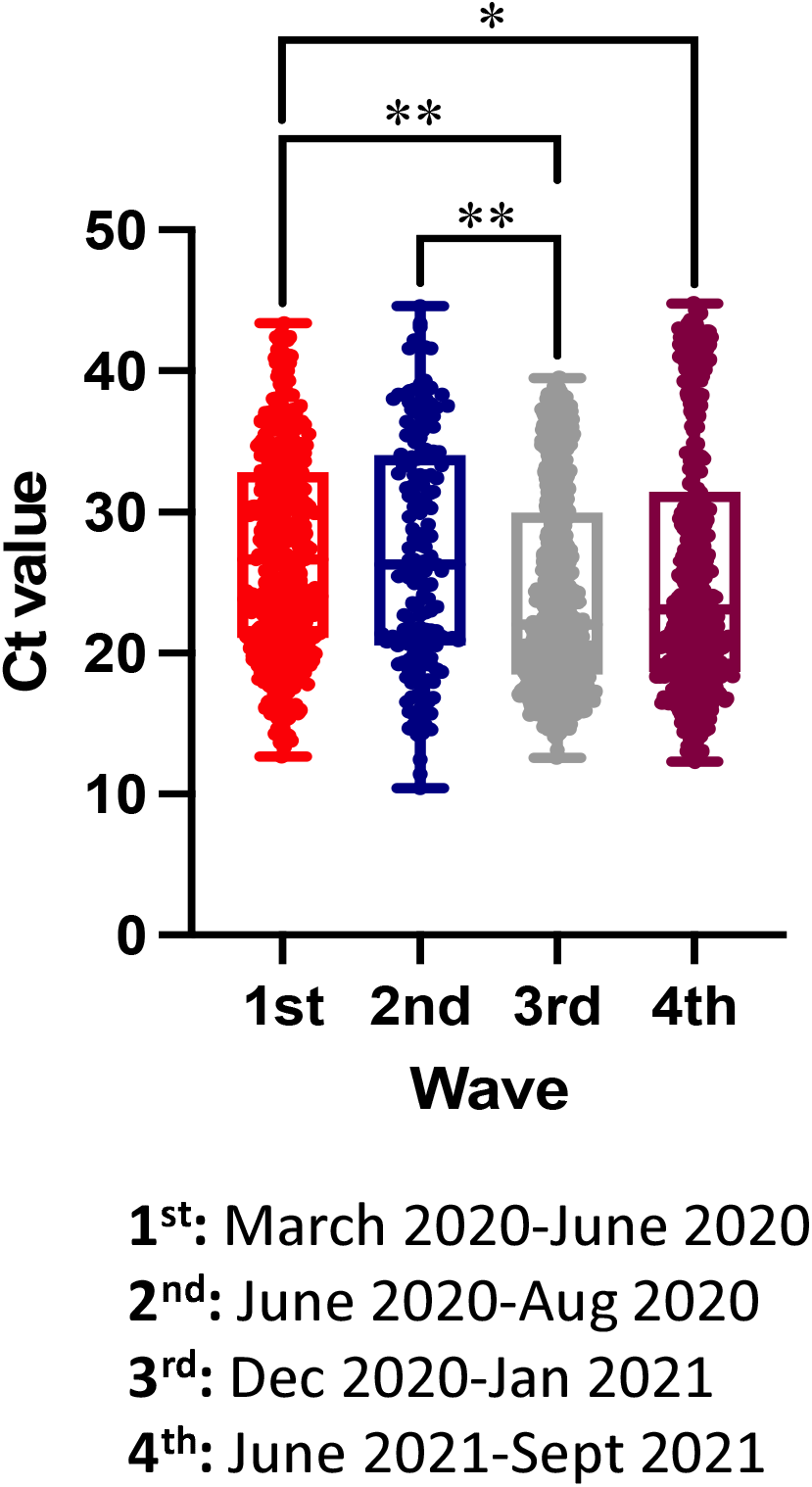
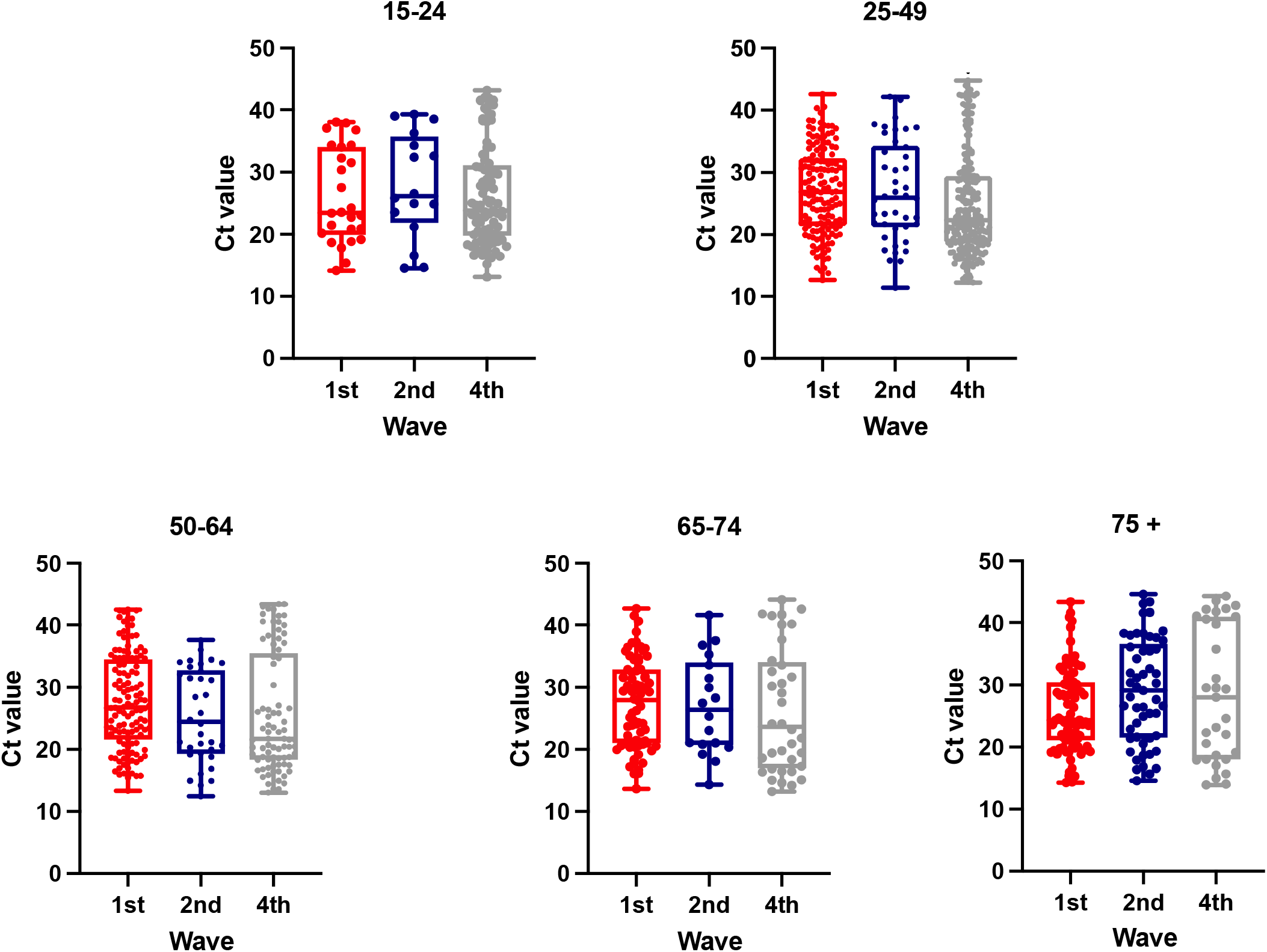
The graphs presented the median cycle threshold (Ct) value of SARS-CoV-2 **A)** in the 1^st^-4^th^ waves of COVID-19 in the UPMC health care system in Allegheny County, Pennsylvania. *p <0.001, **p <0.0001 **B)** in different age groups.

**Figure 2.**
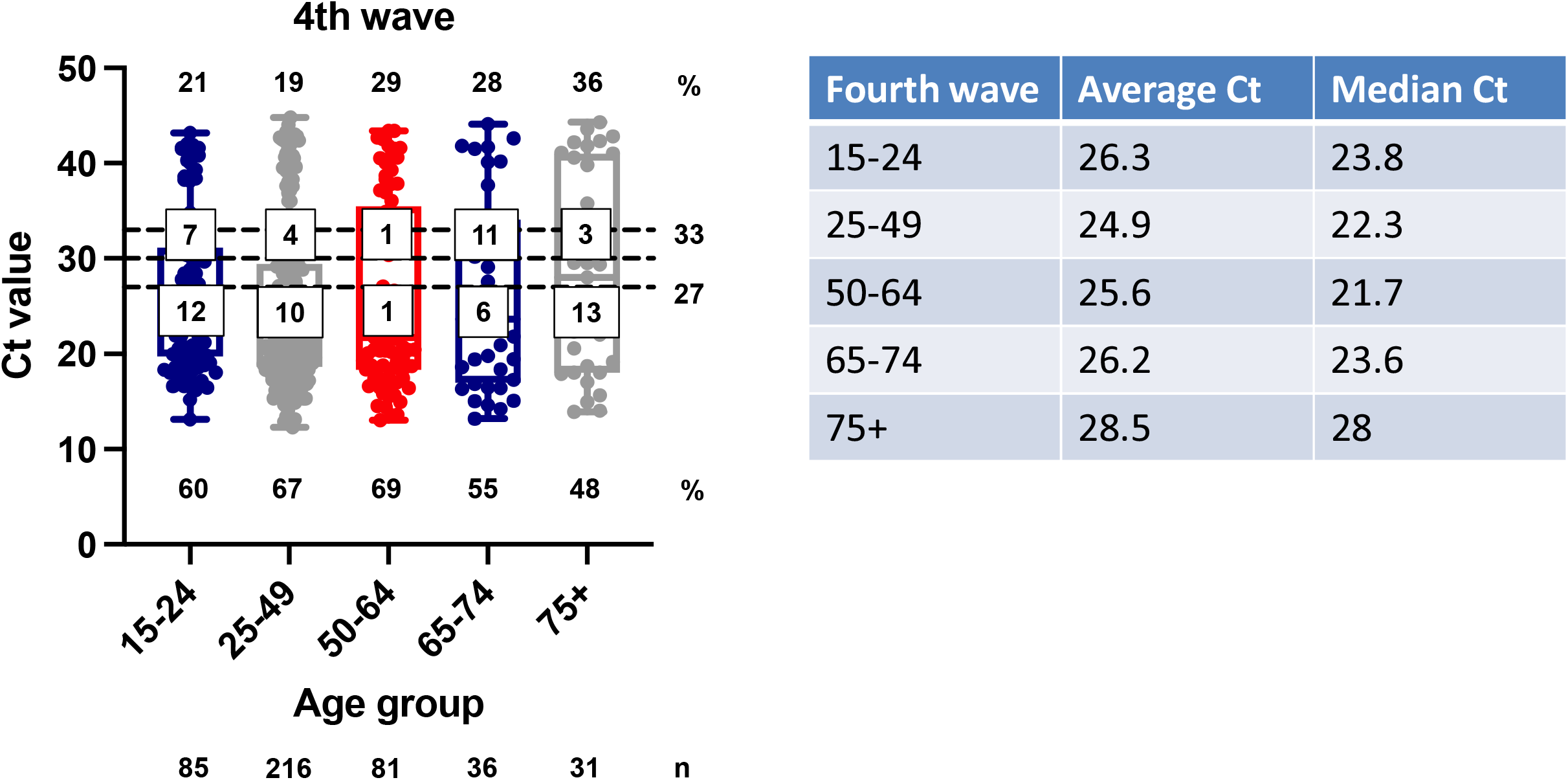
The graph presented the median cycle threshold (Ct) value of SARS-CoV-2 in different age groups in the 4^th^ wave of COVID-19 in the UPMC health care system in Allegheny County, Pennsylvania. The numbers at the bottom of each box and whisker plot is the percentage of population with Ct < 27, on top is the percentage of Ct > 33, and the numbers in the middle are for the percentages between Ct 27-30 and Ct 30-33. The numbers below the graph are the number of positive patients in each.

Viral load in the upper airways is important mainly for detection, as we and others have found little correlation with viral load (Ct) at time of presentation and disease outcome. However, the various rapid antigen assays and some near patient or point of care NAAT technologies detect only to lower Ct values. We, and others have found that antigen tests (RAT) as a group detect nearly all cases that present with Ct of 27 or lower, and that the rapid NAAT such as IDNow is comparable to other NAAT assays at Ct up to 33 (8, 10). To guide our utility of testing, we mapped the detected Ct values in the fourth wave cases against the cutoffs of Ct 27, 30, and 33 (Fig. 2). As can be seen, in the most fragile group of over 75, more 36% would be at risk of being deemed a false negative by the any of the more rapid assays, and less than half (48%) would be consistently detected by RAT assays. Even in the younger age groups, over a third would be missed by RAT assays, and about a quarter by the rapid NAAT assays.

These findings impact our approach to diagnosing SARS-CoV-2 infection. Unfortunately, the somewhat higher viral loads in the upper airways does not translate into a change in our approach of using amplification technologies within the laboratory setting to detect cases in persons who present with symptoms to our hospitals and emergency departments. This is because the risk of missing positive patients in the more vulnerable groups, such as those over 75 years of age, is still quite high. As there are SARS-CoV-2 specific therapies for those with moderate to severe disease, including aggressive use of engineered monoclonal antibodies and remdesivir limited to early in the disease process, correct diagnosis the first time is critical to preventing ventilation and death. For younger, and non-high-risk outpatients not requiring hospitalization or supplemental oxygen such as in healthy children (11), we are using RAT testing with the proviso of missing some quarter of the patients.

## Data Availability

All data produced in the present work are contained in the manuscript

## Acknowledgements

We thank the UPMC Clinical Microbiology Laboratory for testing the specimens.

## Author contributions

TP and AW: designed the study and wrote the manuscript; AM and MM: managed the testing; and JM: performed statistical analyses.

## Conflicts of interest

The authors declare no competing financial interests.

## Ethical approval

All testing was performed as apart of routine clinical care and performed according to CLIA ‘88 regulations by appropriate personnel. The entire study was deemed to be a Quality Improvement initiative by the UPMC IRB and approved by the UPMC QI Review Board.

## Notes

### Competing Interest Statement

The authors have declared no competing interest.

### Funding Statement

This study did not receive any funding

### Author Declarations

Ethics committee/IRB of University of Pittsburgh Medical Center gave ethical approval for this work

## References

1. Cui J, Li F, Shi ZL. 2019. Origin and evolution of pathogenic coronaviruses. Nat Rev Microbiol 17:181–192.

2. Triggle RC, Bansal D, Ding H, Islam MM, Farag E, Hadi AH, Sultan AA. 2021. A comprehensive review of viral characteristics, transmission, pathophysiology, immune response, and management of SARS-CoV-2 and COVID-19 as a basis for controlling the pandemic. Front Immunol 12:631139.

3. Nasserie T, Hittle M, Goodman SN. 2021. Assessment of the frequency and variety of persistent symptoms among patients with COVID-19: a systematic review. JAMA Netw Open 4:e2111417.

4. Ghinai I, McPherson TD, Hunter JC, Kirking HL, Christiansen D, Joshi K, Rubin R, Morales-Estrada S, Black SR, Pacilli M, Fricchione MJ, Chugh RK, Walblay KA, Ahmed NS, Stoecker WC, Hasan NF, Burdsall DP, Reese HE, Wallace M, Wang C, Moeller D, Korpics J, Novosad SA, Benowitz I, Jacobs MW, Dasari VS, Patel MT, Kauerauf J, Charles EM, Ezike NO, Chu V, Midgley CM, Rolfes MA, Gerber SI, Lu X, Lindstrom S, Verani JR, Layden JE; Illinois COVID-19 Investigation Team. 2020. First known person-to-person transmission of severe acute respiratory syndrome coronavirus 2 (SARS-CoV-2) in the USA. Lancet 395:1137–1144.

5. Jarvis CM. 2020. Aerosol transmission of SARS-CoV-2: physical principles and implications. Front Public Health 8:590041.

6. Wang W, Xu Y, Gao R, Lu R, Han K, Wu G, Tan W. 2020. Detection of SARS-CoV-2 in different types of clinical specimens. JAMA 323:1843–1844.

7. Bwire GM, Majigo MV, Njiro BJ, Mawazo A. 2021. Detection profile of SARS-CoV-2 using RT-PCR in different types of clinical specimens: A systematic review and meta-analysis. J Med Virol 93:719–725.

8. Marti JLG, Gribschaw J, McCullough M, Mallon A, Acero J, Kinzler A, Godesky J, Heidenrich K, Iagnemma J, Vanek M, Pasculle AW, Phan T, Hoberman A, Williams VJ, Mitchell S, Wells A. 2021. Differences in detected viral loads guide use of SARS-CoV-2 antigen-detection assays towards symptomatic college students and children. bioRxiv doi:10.1101/2021.01.28.21250365.

9. Mitchell SL, Ventura ES. 2020. Evaluation and comparison of the Hologic Aptima SARS-CoV-2 assay and the CDC 2019-nCoV real-time reverse transcription-PCR diagnostic panel using a four-sample pooling approach. J Clin Microbiol 58:e02241–20.

10. Mitchell SL, St George K. 2020. Evaluation of the COVID19 ID NOW EUA assay. J Clin Virol 128:104429.

11. Shaikh N, Friedlander EJ, Tate PJ, Liu H, Chang CH, Wells A, Hoberman A. 2021. Performance of a rapid SARS-CoV-2 antigen detection assay in symptomatic children. Pediatrics 148:e2021050832.

